# Childhood cognitive ability and self-harm and suicide in later life

**DOI:** 10.1101/2023.02.10.23285747

**Authors:** Matthew H. Iveson, Emily L. Ball, Heather C. Whalley, Ian J. Deary, Simon R. Cox, G. David Batty, Ann John, Andrew M. McIntosh

## Abstract

Self-harm and suicide remain prevalent in later life. For younger adults, work has highlighted an association between higher early-life cognitive ability and lower self-harm and suicide risk. Comparatively little is known about its association with self-harm and suicide among older adults. Furthermore, most work has measured cognitive ability in early adulthood, raising issues of potential confounding by emerging psychiatric conditions.

The present study examined the association between childhood (age 11) cognitive ability and self-harm and suicide risk among a Scotland-wide cohort of older adults (N = 53037), using health data linkage to follow individuals from age 34 to 85. Self-harm events were extracted from hospital admissions and suicide deaths were extracted from national mortality records. Multistate models were used to model transitions between unaffected, self-harm, and then suicide or non-suicide death, and to examine the association between childhood cognitive ability and each transition.

After adjusting for childhood and adulthood socioeconomic conditions, higher childhood cognitive ability was significantly associated with reduced risk of self-harm among older females (N events = 516; HR = 0.90, 95% CI = [0.81, 0.99]). A similar, though non-significant, association was observed among older males (N events = 451; HR = 0.90, 95% CI = [0.82, 1.00]). Although suicide risk was higher among older adults experiencing self-harm, childhood cognitive ability was not significantly associated with suicide risk among either older adults experiencing no self-harm events (Male: N events = 118, HR = 1.17, 95% CI = [0.84, 1.63]; Female: N events = 31, HR = 1.30, 95% CI = [0.70, 2.41]) or those experiencing a self-harm event during follow-up (Male: N events = 16, HR = 1.05, 95% CI = [0.61, 1.80]; Female: N events = 13, HR = 1.08, 95% CI = [0.55, 2.14]). Higher suicide risk was significantly associated with covariates including higher adulthood deprivation and longer time in the self-harm state. These results extend work on cognitive ability and mental health, demonstrating that these associations can span across the life course and into older age.

## Introduction

Self-harm and suicide are common among middle-aged and older adults. A recent meta-analysis reported yearly rates of between 19 and 65 per 100,000 in older adults (aged 60+) for self-harm (Troya et al., 2019) and recent UK estimates show completed suicide rates peak in middle-age (England and Wales: 15 per 100,000; Scotland: 21 per 100,000) and drop only slightly (England & Wales: 10 per 100,000; Scotland: 12 per 100,000) after the age of 60 (ONS, 2021; Scottish Public Health Observatory, 2022).

A number of psychosocial risk factors have been identified for self-harm and suicide, including socioeconomic disadvantage, social isolation, chronic distress and psychiatric disorders (Batty et al., 2018). Among such risk factors, cognitive ability (intelligence) has been associated with both self-harm and suicide risk. Evidence from large cohort studies suggests that higher cognitive ability is associated with lower risk of self-harm (e.g., HR = 0.64 per standard deviation increase in cognitive ability) (Batty et al., 2010; Fergusson et al., 2005; Jiang et al., 1999; Osler et al., 2008) and lower risk of suicide (e.g., HR = 0.70 per standard deviation increase in cognitive ability) (Allebeck et al., 1988, p. 988; Andersson et al., 2008; Christensen et al., 2016; Gunnell et al., 2005; Hemmingsson et al., 2006; Osler et al., 2008; O’Toole & Cantor, 1995). However, the majority of these studies focus on self-harm and suicide risk up to middle-age, particularly among males. The epidemiology of self-harm and suicide may change in later life, and it is unclear whether cognitive ability also predicts self-harm and suicide risk into older age, for both males and females. Furthermore, the majority of studies use cognitive ability measured in early-adulthood; using a measure from childhood may minimise the impact of confounders such as education and undetected psychiatric conditions (Hatch et al., 2007; Kuh et al., 2004).

Only one study has examined the association between childhood cognitive ability and suicide into later-life for both males and females. In a nationwide sample of Scottish older adults, Calvin et al. (2017) reported that higher childhood cognitive ability was associated with a reduced risk of suicide (from age 11 to age 79) among males (HR = 0.80, 95% CI [0.66, 0.96]), but not among females (HR = 1.15, 95% CI [0.82, 1.60]). No studies to date have examined the impact of childhood cognitive ability on self-harm in older adults. Although self-harm and suicide share common risk factors (Duarte et al., 2020), it is unclear whether childhood cognitive ability is a risk factor for suicide specifically, or whether its contribution may be mediated by self-harm.

The majority of work – including that in later-life (Calvin et al., 2017) – does not consider both self-harm and suicide concurrently. This makes it difficult to estimate the independent contribution of cognitive ability. Although self-harm and suicide are distinct outcomes, self-harm is one of the most important risk factors for suicide, present in almost half of those who go on to take their own lives (Chan et al., 2016; Hawton et al., 2003). Indeed, the risk of suicide among older adults presenting to hospital with self-harm has been shown to be 67 times higher than the general population of older adults (Murphy et al., 2012). Self-harm may act as a mediator of the association between cognitive ability and suicide risk. Examining self-harm and suicide concurrently is important for disentangling risk, and for estimating the association between childhood cognitive ability and suicide separately among those with and without self-harm behaviours.

The present study examines the association between childhood cognitive ability and later-life self-harm and suicide in a population birth cohort recruited in childhood. After follow-up for suicide risk in the Scottish Mental Survey 1947 cohort (Calvin et al., 2017), the present study uses multistate models to examine the independent, concurrent associations between childhood cognitive ability and transitions into both self-harm and suicide. The present study also extends the follow-up period to cover routinely-collected health records from middle-age up to age 85.

## Methods

### Sample

The sample selection process is shown in Figure 1. The initial sample was formed of 70,805 individuals who took part in the Scottish Mental Survey 1947 (SMS1947) (The Scottish Council for Research in Education, 1949), a nationwide assessment of cognitive ability and socioeconomic conditions administered to 1936-born individuals attending a school in Scotland in June 1947. The analytic sample was formed of 53,037 of these individuals who were traceable in Community Health Index (CHI) records. A CHI record is generated upon an individual’s first contact with the National Health Service (NHS) in Scotland; individuals are assigned a unique number that is present on most electronic health records enabling them to be linked together. Excluded participants (N = 19,104) either could not be traced in CHI-linked records or had multiple potential matches without a means to reconcile them. De-identified datasets containing person-level data were created by Public Health Scotland Records and were made accessible to named researchers within a secure Trusted Research Environment.

**Figure 1.**
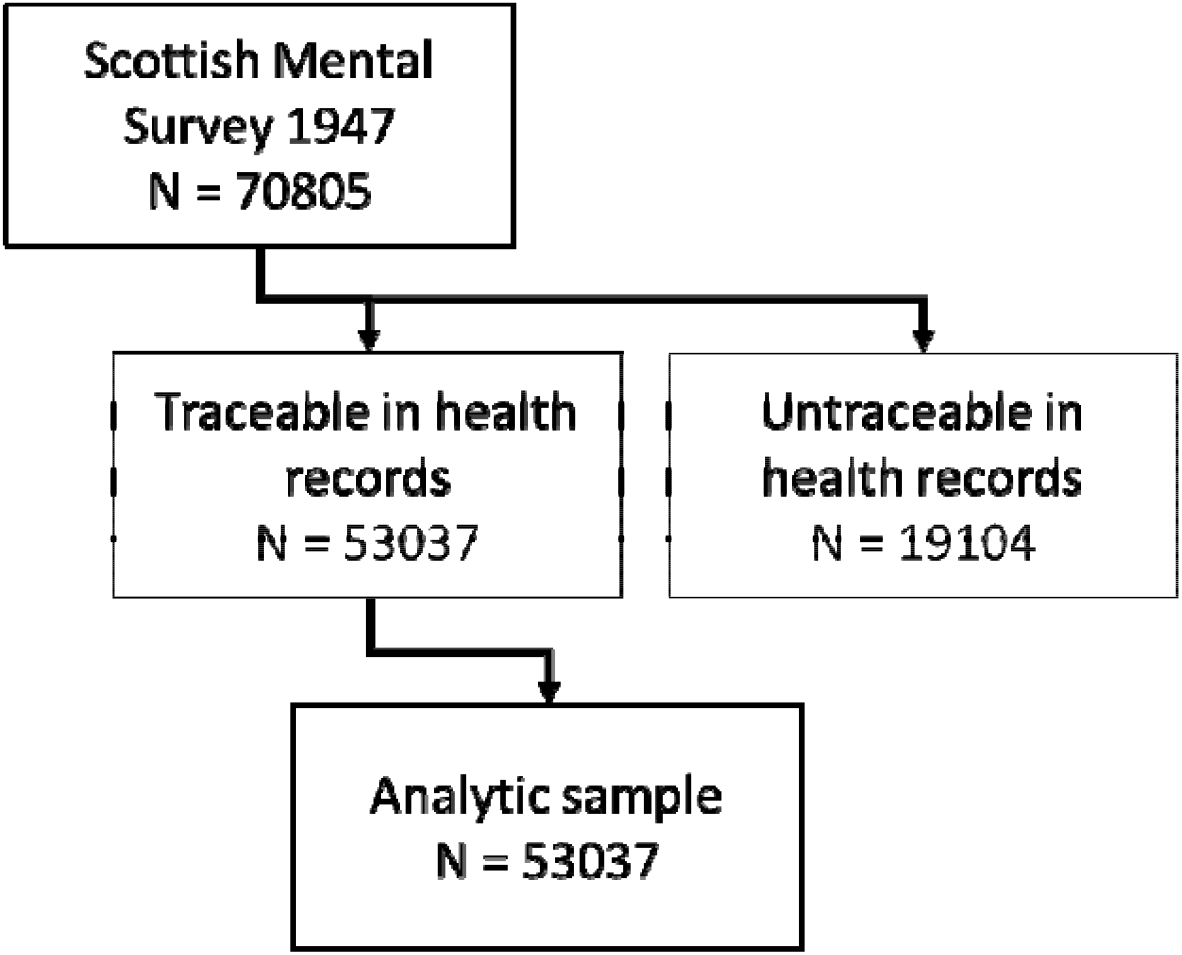
Sample selection

### Self-harm-related hospitalisation, suicide and other deaths

Self-harm and suicide outcomes were extracted from routinely-collected national health records. Self-harm-related hospitalisations were taken from Scottish Morbidity Records (SMR), covering all inpatient admissions to a Scottish NHS hospital from January 1980 to March 2021 (age 44 to age 85 years-old). General/acute inpatient and day case (SMR01) and mental health inpatient and day case (SMR04) admissions were examined for diagnoses that matched a list of ICD9 and ICD10 codes related to intentional self-injury and poisonings (Supplementary Material). Note that this definition does not distinguish between attempted suicide and other reasons for self-harm. However, attempted suicide makes up a greater proportion of self-harm presentations to hospital, particularly among older adults (Draper, 1996; Hawton & Harriss, 2008). The definition of self-harm events in the present sample, then, may predominantly represent suicide attempts. A binary variable for each admission was created indicating a self-harm-related hospital admission. Finally, a binary variable was created to indicate whether an individual had experienced a self-harm-related hospitalisation and the age (in months) at which the first self-harm admission occurred.

Deaths due to suicide and other causes of death were taken from the National Records of Scotland Death Register, covering all deaths registered in Scotland from January 1970 to March 2021 (age 34 to age 85 years-old). Deaths due to intentional self-injury and poisonings are flagged within the register by the data provider using a list of ICD9 and ICD10 codes (X60-X84). Note that many previous studies – including population estimates by the ONS (ONS, 2021) – use a broader classification that includes deaths due to undetermined events. In Scotland, these are operationalised as ‘probable self-harm’ and so are not included in the present definition of suicide. However, self-injury and undetermined (open) verdicts have been shown to be largely similar in terms of their demographic and medical parameters (Linsley et al., 2001). The flag in death records was used to create a binary variable indicating whether a death had been due to suicide or due to other causes. Age (in months) at which the death occurred was also recorded.

### Childhood cognitive ability

Cognitive ability in childhood was measured using the Moray House Test No. 12 (MHT), completed on the 4^th^ of June 1947, around age 11, as part of the SMS1947 (The Scottish Council for Research in Education, 1949). The MHT is a group-administered assessment of general cognitive ability, including items on verbal reasoning, arithmetic and spatial problem solving. Participants could score a maximum of 76. Raw scores were age-residualised to account for small variations in age at test, before being IQ-scaled (M = 100, SD = 15) and standardised (M = 0, SD = 1) to aid interpretation.

### Covariates

Sex (male or female) was taken from routinely-collected NHS CHI records. Childhood conditions were measured using a two-item survey completed as part of the SMS1947. Participants were asked to report the number of children present in their family (family size) and their position among children ordered by age (family position). Higher numbers in family size indicated larger families, and higher numbers in family position indicated younger age relative to other children in the family. Family size was Yeo-Johnson scaled (a monotonic transformation using power functions) to account for a non-normal distribution.

Socioeconomic conditions in adulthood were measured by the decile on the Carstairs Index 1991, an area-based measure of relative material deprivation that aggregates male unemployment, low occupational social class, non-car ownership and household overcrowding over a data zone (Carstairs & Morris, 1991; McLoone, 2004). Carstairs deciles are recorded routinely as part of admission to secondary care NHS services in Scotland, including in those records linked as part of the present study. For those experiencing self-harm, deciles were taken from the hospital inpatient record associated with a first self-harm admission. For those not experiencing self-harm, deciles were taken from their last recorded inpatient admission (i.e., closest to death, suicide or censoring at end of follow-up). Those without any inpatient admissions were treated as having missing Carstairs 1991 deciles.

### Statistical analyses

Analyses were conducted for males and females separately due to reported sex differences in both self-harm and suicide (Chan et al., 2007; Murphy et al., 2012) and mortality (Nathanson, 1984; Read & Gorman, 2010) risk. Multistate models were used to account for the fact that individuals could move between states of health (unaffected at baseline), self-harm, and suicide and other causes of death in several ways. Figure 2 models the possible transitions. All individuals entered the study as unaffected, though this does not account for health (including previous episodes of self-harm) prior to entry. Notably, self-harm, suicide and other causes of death were treated as absorbing states, with no return to the unaffected state allowed. Furthermore, suicide and other causes of death were treated as competing states; individuals could only enter one of these states with no transition between them. Time to transition was estimated in months from birth. Individuals were right censored at 1027 months (around 86 years-old) at the end of the follow-up period.

**Figure 2.**
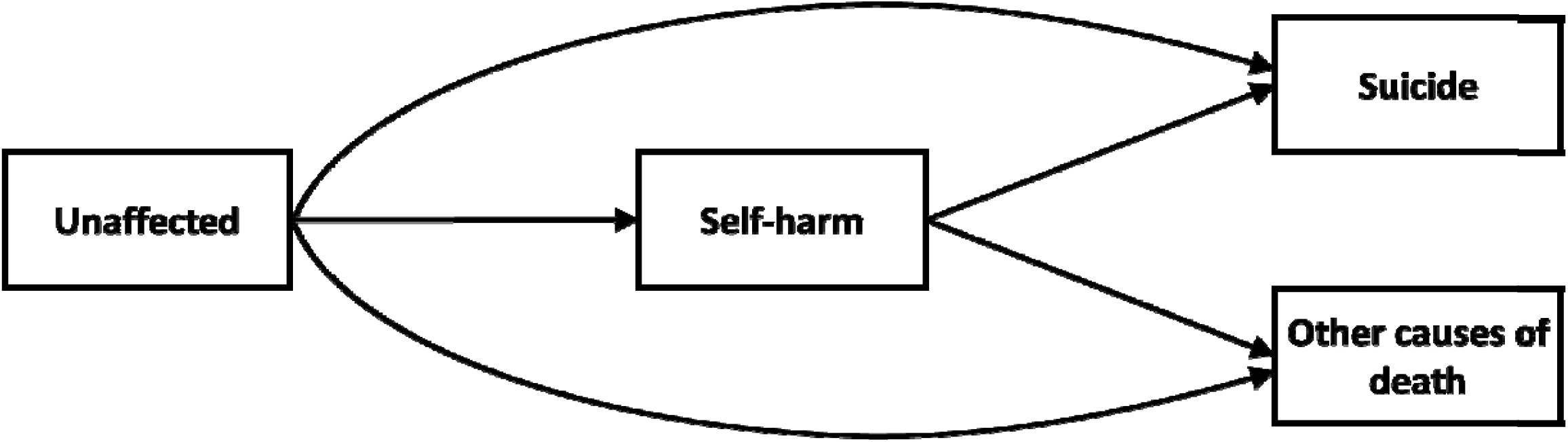
Transition diagram indicating five possible transitions modelled in the analyses. All individuals enter the study in the unaffected state

A univariate model was constructed to assess the association between childhood cognitive ability and the hazards of each of the 5 transitions (Figure 2) simultaneously. Similar univariate models were created for each of the covariates and their associations with the 5 transitions. A multivariate model was then conducted including childhood cognitive ability and covariates (sex, family size, family position, Carstairs decile at admission), as well as time (in months) spent in the self-harm state (i.e., time from a self-harm admission to entry to another state). Note that time in the self-harm state associations were only estimated with transitions from self-harm to suicide or to other death.

Analyses were conducted with R (version 4.0.5) (R Core Team, 2021) using the ‘mstate’ package (version 0.3.2) (de Wreede et al., 2011).

## Results

In terms of sample selection effects, individuals excluded due to missing CHI records did not significantly differ from those retained in the analytic sample in age (in months) at SMS1947 (excluded: M = 130.75, SD = 3.44; analytic sample: M = 130.75, SD = 3.44; t = -0.18, df = 33823, p = 0.85). However, excluded participants had significantly higher MHT scores (excluded: M = 38.47, SD = 15.76; analytic sample: M = 36.39, SD = 15.71; t = 15.12, df = 31460, p < 0.001), significantly earlier position in family (excluded: M = 2.45, SD = 1.77; analytic sample: M = 2.53, SD = 1.82; t = -5.11, df = 34344, p < 0.001) and significantly smaller families (excluded: M = 3.70, SD = 2.16; analytic sample: M = 3.80, SD = 2.22; t = -5.46, df = 34328, p < 0.001) at age 11.

Descriptive statistics for the analytic sample are shown in Table 1. Of the 53,037 individuals in the analytic sample, 28,097 appeared in death records (N = 178 with a ‘suicide’ cause of death) and 16,895 appeared in hospital inpatient records (N = 967 with a self-harm-related admission). There was no significant difference in the proportion of individuals experiencing self-harm between males and females (X^2^ = 3.14, p = 0.08). However, there were significant demographic differences to justify splitting further analyses by sex: compared to males, females exhibited significantly higher MHT scores at age 11 (mean difference = 2.02, p < 0.001), significantly older age at admission (mean difference = 33.05 months, p < 0.001) and significantly older age at censor (mean difference = 26.59 months, p < 0.001). Additionally, a greater proportion of females were alive at the end of follow-up relative to males (X^2^ = 1024.53, p < 0.001).

**Table 1.** Descriptive statistics for the analytic sample according to sex

|  |  | **Female**<br>**N = 26803** | **Male**<br>**N = 26234** | **Total**<br>**N = 53037** |
| --- | --- | --- | --- | --- |
| **Age 11 MHT score** |  |  |  |  |
|  | Mean (SD) | 37.39<br>(14.98) | 35.37<br>(16.36) | 36.39<br>(15.71) |
|  | N Missing (%) | 1773<br>(7) | 1753<br>(7) | 3526<br>(7) |
| **Age 11 position in family** |  |  |  |  |
|  | Mean (SD) | 2.55<br>(1.85) | 2.51<br>(1.80) | 2.53<br>(1.82) |
|  | N Missing (%) | 269<br>(1) | 226<br>(1) | 495<br>(1) |
| **Age 11 size of family** |  |  |  |  |
|  | Mean (SD) | 3.83<br>(2.25) | 3.78<br>(2.20) | 3.80<br>(2.22) |
|  | N Missing (%) | 272 | 227 | 499 |
| **Carstairs decile at admission** |  |  |  |  |
|  | Median (IQR) | 6<br>(5) | 6<br>(5) | 6<br>(5) |
|  | N Missing (%) | 17789<br>(66) | 18242<br>(70) | 36031<br>(68) |
| **Age at admission (months)** |  |  |  |  |
|  | Mean (SD) | 865.12<br>(126.49) | 832.07<br>(134.07) | 849.57<br>(131.15) |
|  | N Missing (%) | 17729<br>(66) | 18175<br>(69) | 35904<br>(68) |
| **Age at censor (months)** |  |  |  |  |
|  | Mean (SD) | 929.66<br>(119.27) | 903.07<br>(128.63) | 916.51<br>(124.70) |
|  | N Missing (%) | 0<br>(0) | 0<br>(0) | 0<br>(0) |
| **Self-harm status** |  |  |  |  |
|  | N No self-harm (%) | 26287<br>(98) | 25783<br>(98) | 52070<br>(98) |
|  | N Self-harm (%) | 516<br>(2) | 451<br>(2) | 967<br>(2) |
|  | N Missing (%) | 0<br>(0) | 0<br>(0) | 0<br>(0) |
| **Deceased status** |  |  |  |  |
|  | N Alive (%) | 14419<br>(54) | 10521<br>(40) | 24940<br>(47) |
|  | N Suicide (%) | 44 (<1) | 134 (<1) | 178 (<1) |
|  | N Other Deceased (%) | 12340<br>(46) | 15579<br>(60) | 27919<br>(53) |
|  | N Missing (%) | 0 | 0 | 0 |
|  |  | (0) | (0) | (0) |

MHT = Moray House Test measuring cognitive ability; Carstairs = Carstairs Index 1991 measuring area-level deprivation

Transitions between states are shown in Tables 2 (males) and 3 (females). Kaplan-Meier curves showing cumulative hazards for each transition are shown in Figure 3. Transitions to non-suicide death were more frequent among males (Cumulative Incidence = 59.38%) than females (Cumulative Incidence = 46.04%). In contrast, no transition (i.e., no self-harm, suicide or death) was more frequent among females (Cumulative Incidence = 53.23%) than males (Cumulative Incidence = 39.78%). Transitions to self-harm were slightly more frequent among females (Cumulative Incidence = 1.93%) than males (Cumulative Incidence = 1.72%). Examining the relative risk of suicide among those with and without self-harm events (hospital admissions) demonstrated a much higher ratio among females (RR = 21.00, 95%CI [11.47, 41.38]) than among males (RR = 7.89, 95%CI [4.72, 13.18]).

**Table 2.** Frequency of each transition, Males (N = 26234). Diagonals represent those who stayed in each state until the end of follow-up

|  | To |  |  |  |  |
| --- | --- | --- | --- | --- | --- |
| From |  | Unaffected | Self-harm | Suicide | Death |
|  | Unaffected | 10435 | 451 | 118 | 15230 |
|  | Self-harm | - | 86 | 16 | 349 |
|  | Suicide | - | - | 134 | - |
|  | Death | - | - | - | 15579 |

**Table 3.** Frequency of each transition, Females (N = 26803). Diagonals represent those who stayed in each state until end of follow-up

|  | To |  |  |  |  |
| --- | --- | --- | --- | --- | --- |
| From |  | Unaffected | Self-harm | Suicide | Death |
|  | Unaffected | 14266 | 516 | 31 | 11990 |
|  | Self-harm | - | 153 | 13 | 350 |
|  | Suicide | - | - | 44 | - |
|  | Death | - | - | - | 12340 |

**Figure 3.**
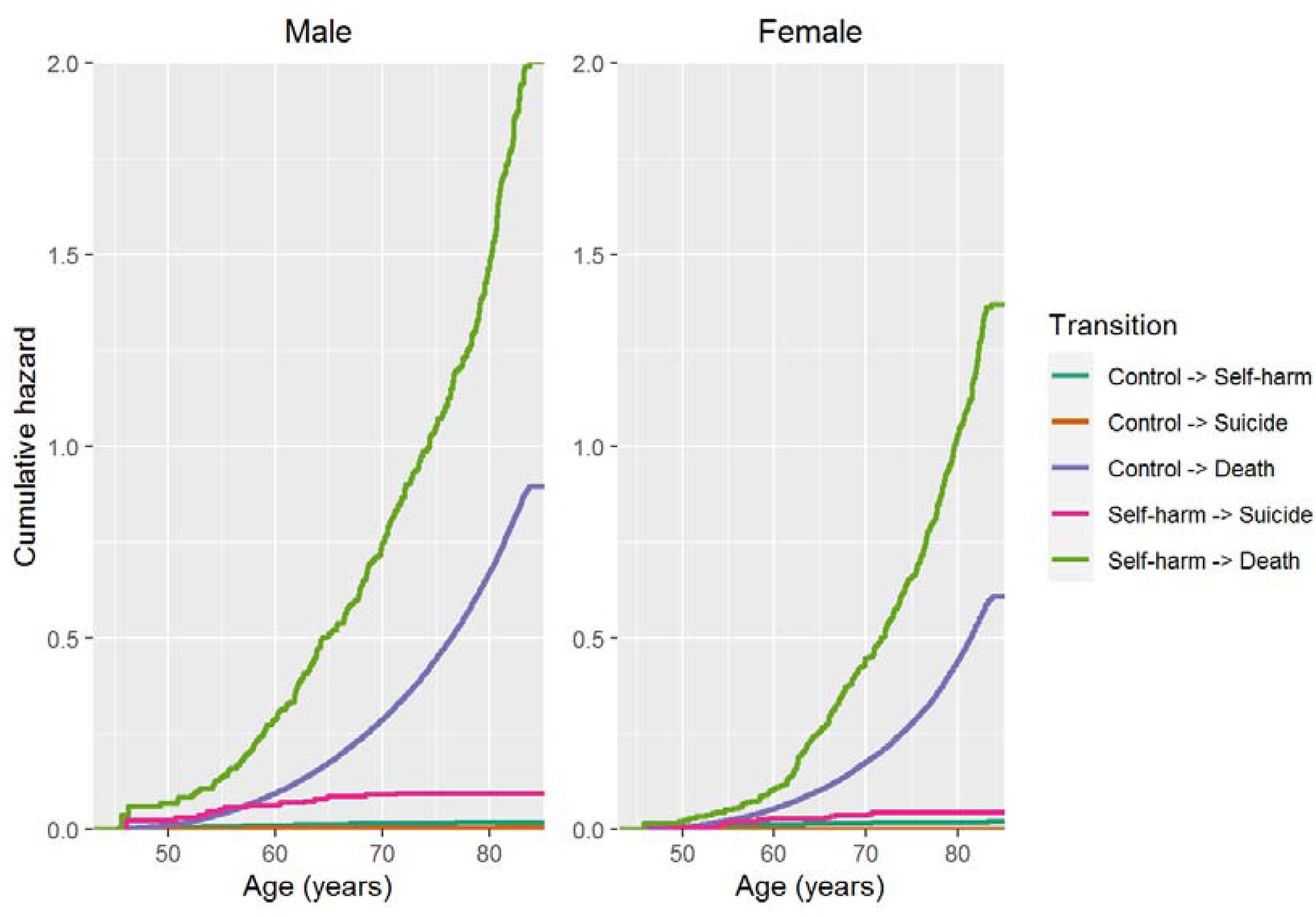
Kaplan-Meier curves showing cumulative hazards for each transition, for males and females

### Self-harm risk

The univariate and fully-adjusted associations between each variable and the risk of each transition are shown in Table 4 (Males) and Table 5 (Females). Lower childhood cognitive ability was significantly associated with an increased risk of transitioning from unaffected to self-harm states, though this only remained significant in the mutually-adjusted model among females. In females, a 1SD advantage in childhood cognitive ability was associated with a 10% lower risk of self-harm (HR = 0.90, 95% CI [0.81, 0.99]). Although non-significant, a similar strength of association was observed among males (HR = 0.90, 95% CI [0.82, 1.00]). Re-running the mutually-adjusted model with a combined sample of males and females showed a significant main effect of childhood cognitive ability (HR = 0.88, 95% CI [0.79, 0.96], p = 0.01), but no significant main effect of sex (HR = 1.02, 95% CI [0.89, 1.17], p = 0.80) and no significant interaction between sex and childhood cognitive ability (HR = 1.06, 95% CI [0.93, 1.20], p = 0.42). This indicates apparent sex-differences in the association between childhood cognitive ability and self-harm risk are likely due to differences in statistical power. Higher adult deprivation was significantly associated with increased risk of self-harm transitions among both males and females, including in the mutually-adjusted model. A single decile increase on the Carstairs index of deprivation (indicating more deprivation) was associated with a 7% higher risk of transitioning to self-harm among males (HR = 1.07, 95% CI [1.03, 1.11]) and a 12% increase in risk among females (HR = 1.12, 95% CI [1.08, 1.16]).

**Table 4.**
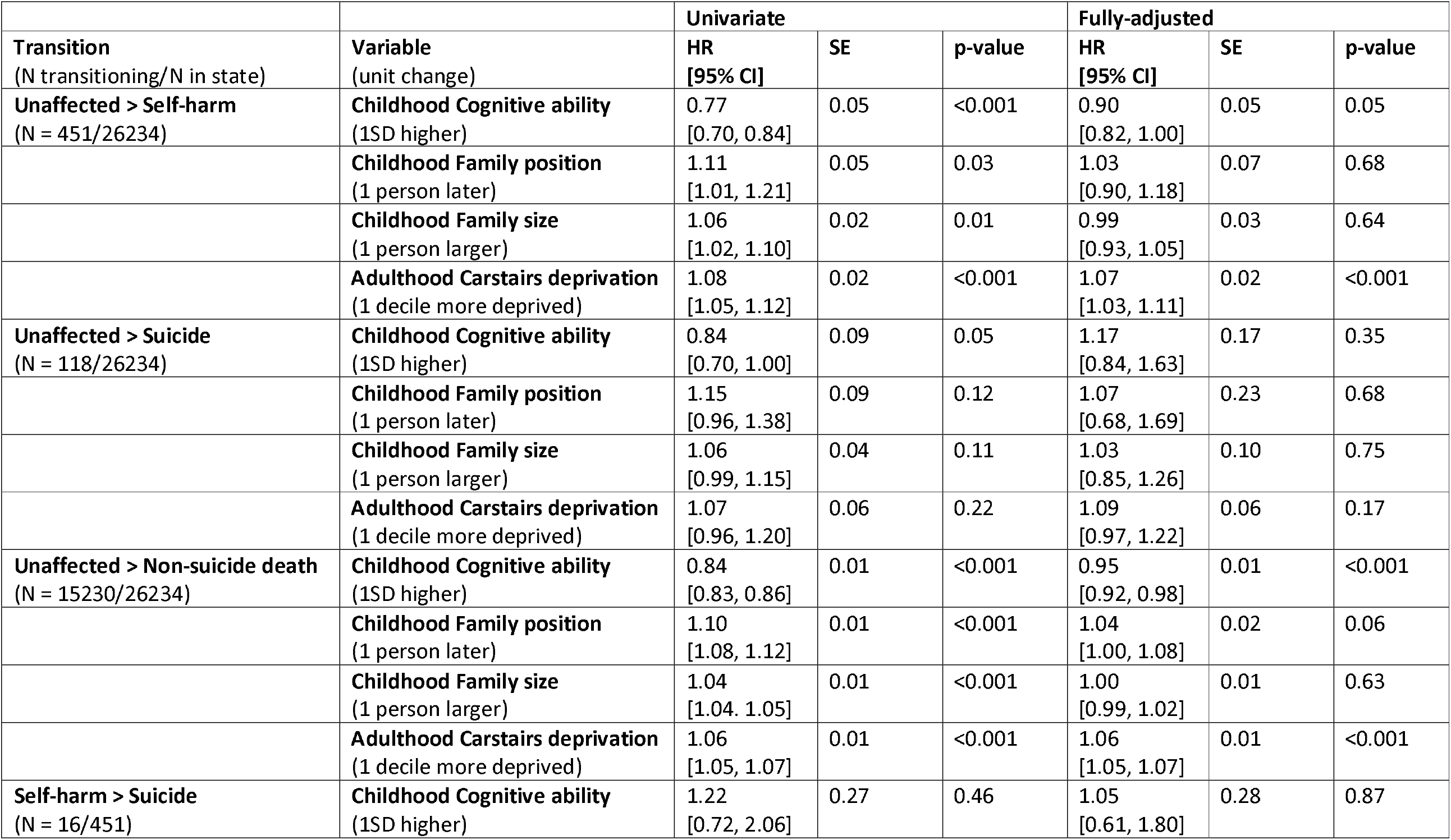
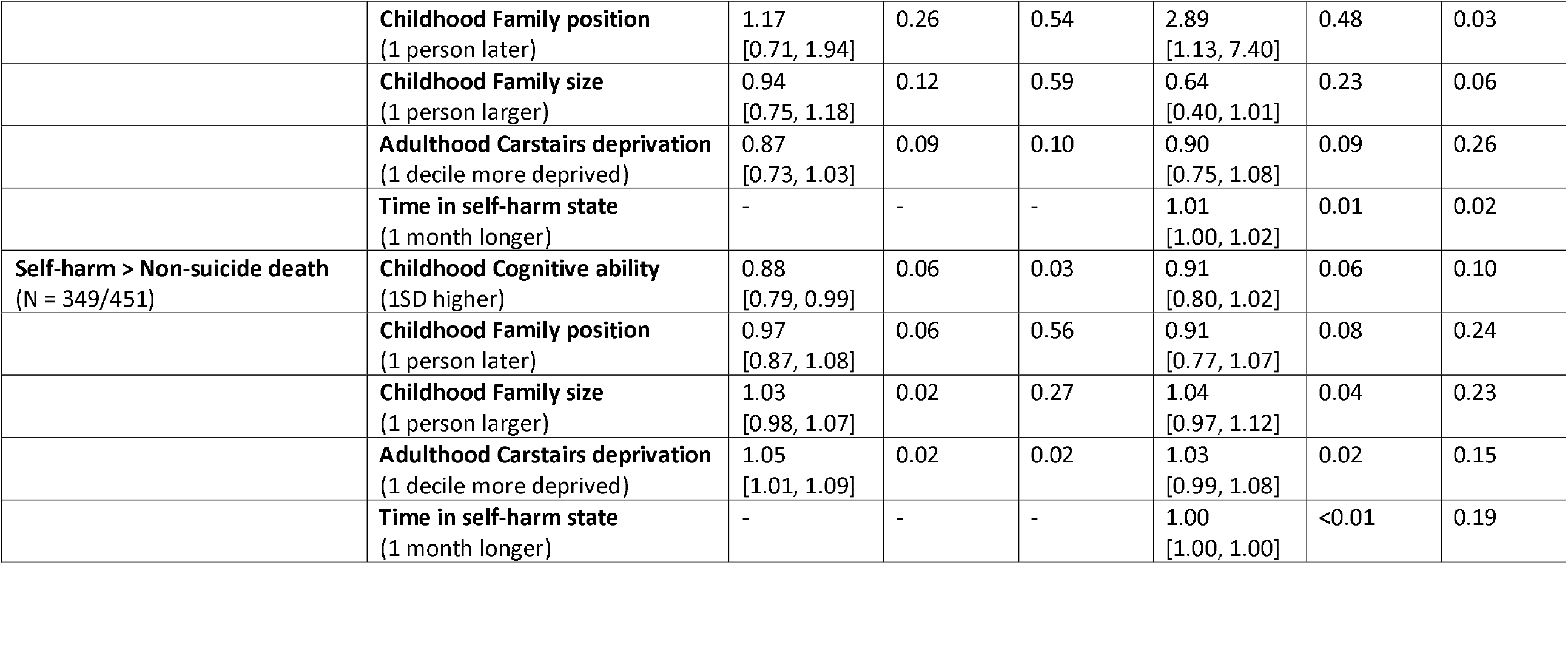
Univariate and fully-adjusted associations with each transition from the multistate model, males

| Transition<br>(N transitioning/N in state) | Variable<br>(unit change) | Univariate |  |  | Fully-adjusted |  |  |
| --- | --- | --- | --- | --- | --- | --- | --- |
|  |  | HR<br>[95% CI] | SE | p-value | HR<br>[95% CI] | SE | p-value |
| **Unaffected > Self-harm**<br>(N = 451/26234) | **Childhood Cognitive ability**<br>(1SD higher) | 0.77<br>[0.70, 0.84] | 0.05 | <0.001 | 0.90<br>[0.82, 1.00] | 0.05 | 0.05 |
|  | **Childhood Family position**<br>(1 person later) | 1.11<br>[1.01, 1.21] | 0.05 | 0.03 | 1.03<br>[0.90, 1.18] | 0.07 | 0.68 |
|  | **Childhood Family size**<br>(1 person larger) | 1.06<br>[1.02, 1.10] | 0.02 | 0.01 | 0.99<br>[0.93, 1.05] | 0.03 | 0.64 |
|  | **Adulthood Carstairs deprivation**<br>(1 decile more deprived) | 1.08<br>[1.05, 1.12] | 0.02 | <0.001 | 1.07<br>[1.03, 1.11] | 0.02 | <0.001 |
| **Unaffected > Suicide**<br>(N = 118/26234) | **Childhood Cognitive ability**<br>(1SD higher) | 0.84<br>[0.70, 1.00] | 0.09 | 0.05 | 1.17<br>[0.84, 1.63] | 0.17 | 0.35 |
|  | **Childhood Family position**<br>(1 person later) | 1.15<br>[0.96, 1.38] | 0.09 | 0.12 | 1.07<br>[0.68, 1.69] | 0.23 | 0.68 |
|  | **Childhood Family size**<br>(1 person larger) | 1.06<br>[0.99, 1.15] | 0.04 | 0.11 | 1.03<br>[0.85, 1.26] | 0.10 | 0.75 |
|  | **Adulthood Carstairs deprivation**<br>(1 decile more deprived) | 1.07<br>[0.96, 1.20] | 0.06 | 0.22 | 1.09<br>[0.97, 1.22] | 0.06 | 0.17 |
| **Unaffected > Non-suicide death**<br>(N = 15230/26234) | **Childhood Cognitive ability**<br>(1SD higher) | 0.84<br>[0.83, 0.86] | 0.01 | <0.001 | 0.95<br>[0.92, 0.98] | 0.01 | <0.001 |
|  | **Childhood Family position**<br>(1 person later) | 1.10<br>[1.08, 1.12] | 0.01 | <0.001 | 1.04<br>[1.00, 1.08] | 0.02 | 0.06 |
|  | **Childhood Family size**<br>(1 person larger) | 1.04<br>[1.04, 1.05] | 0.01 | <0.001 | 1.00<br>[0.99, 1.02] | 0.01 | 0.63 |
|  | **Adulthood Carstairs deprivation**<br>(1 decile more deprived) | 1.06<br>[1.05, 1.07] | 0.01 | <0.001 | 1.06<br>[1.05, 1.07] | 0.01 | <0.001 |
| **Self-harm > Suicide**<br>(N = 16/451) | **Childhood Cognitive ability**<br>(1SD higher) | 1.22<br>[0.72, 2.06] | 0.27 | 0.46 | 1.05<br>[0.61, 1.80] | 0.28 | 0.87 |
|  | **Childhood Family position**<br>(1 person later) | 1.17<br>[0.71, 1.94] | 0.26 | 0.54 | 2.89<br>[1.13, 7.40] | 0.48 | 0.03 |
|  | **Childhood Family size**<br>(1 person larger) | 0.94<br>[0.75, 1.18] | 0.12 | 0.59 | 0.64<br>[0.40, 1.01] | 0.23 | 0.06 |
|  | **Adulthood Carstairs deprivation**<br>(1 decile more deprived) | 0.87<br>[0.73, 1.03] | 0.09 | 0.10 | 0.90<br>[0.75, 1.08] | 0.09 | 0.26 |
|  | **Time in self-harm state**<br>(1 month longer) | - | - | - | 1.01<br>[1.00, 1.02] | 0.01 | 0.02 |
| **Self-harm > Non-suicide death**<br>(N = 349/451) | **Childhood Cognitive ability**<br>(1SD higher) | 0.88<br>[0.79, 0.99] | 0.06 | 0.03 | 0.91<br>[0.80, 1.02] | 0.06 | 0.10 |
|  | **Childhood Family position**<br>(1 person later) | 0.97<br>[0.87, 1.08] | 0.06 | 0.56 | 0.91<br>[0.77, 1.07] | 0.08 | 0.24 |
|  | **Childhood Family size**<br>(1 person larger) | 1.03<br>[0.98, 1.07] | 0.02 | 0.27 | 1.04<br>[0.97, 1.12] | 0.04 | 0.23 |
|  | **Adulthood Carstairs deprivation**<br>(1 decile more deprived) | 1.05<br>[1.01, 1.09] | 0.02 | 0.02 | 1.03<br>[0.99, 1.08] | 0.02 | 0.15 |
|  | **Time in self-harm state**<br>(1 month longer) | - | - | - | 1.00<br>[1.00, 1.00] | <0.01 | 0.19 |

**Table 5.** Univariate and fully-adjusted associations with each transition from the multistate model, females

| Transition<br>(N transitioning/N in state) | Variable<br>(unit change) | Univariate |  |  | Fully-adjusted |  |  |
| --- | --- | --- | --- | --- | --- | --- | --- |
|  |  | HR<br>[95% CI] | SE | p-value | HR<br>[95% CI] | SE | p-value |
| **Unaffected > Self-harm**<br>(N = 516/26803) | **Childhood Cognitive ability**<br>(1SD higher) | 0.75<br>[0.68, 0.82] | 0.05 | <0.001 | 0.90<br>[0.81, 0.99] | 0.05 | 0.03 |
|  | **Childhood Family position**<br>(1 person later) | 1.13<br>[1.03, 1.23] | 0.04 | 0.01 | 1.02<br>[0.90, 1.16] | 0.07 | 0.77 |
|  | **Childhood Family size**<br>(1 person larger) | 1.06<br>[1.02, 1.10] | 0.02 | <0.001 | 1.00<br>[0.95, 1.06] | 0.03 | 0.91 |
|  | **Adulthood Carstairs deprivation**<br>(1 decile more deprived) | 1.13<br>[1.10, 1.17] | 0.02 | <0.001 | 1.12<br>[1.08, 1.16] | 0.02 | <0.001 |
| **Unaffected > Suicide**<br>(N = 31/26803) | **Childhood Cognitive ability**<br>(1SD higher) | 1.40<br>[0.92, 2.13] | 0.22 | 0.12 | 1.30<br>[0.70, 2.41] | 0.31 | 0.40 |
|  | **Childhood Family position**<br>(1 person later) | 1.03<br>[0.73, 1.46] | 0.18 | 0.86 | 1.47<br>[0.65, 3.33] | 0.42 | 0.36 |
|  | **Childhood Family size**<br>(1 person larger) | 0.95<br>[0.80, 1.12] | 0.09 | 0.52 | 0.79<br>[0.52, 1.19] | 0.21 | 0.26 |
|  | **Adulthood Carstairs deprivation**<br>(1 decile more deprived) | 1.02<br>[0.85, 1.22] | 0.09 | 0.86 | 1.10<br>[0.91, 1.34] | 0.10 | 0.33 |
| **Unaffected > Non-suicide death**<br>(N = 11990/26803) | **Childhood Cognitive ability**<br>(1SD higher) | 0.79<br>[0.77, 0.80] | 0.01 | <0.001 | 0.84<br>[0.81, 0.87] | 0.02 | <0.001 |
|  | **Childhood Family position**<br>(1 person later) | 1.08<br>[1.06, 1.10] | 0.01 | <0.001 | 0.97<br>[0.93, 1.02] | 0.02 | 0.23 |
|  | **Childhood Family size**<br>(1 person larger) | 1.04<br>[1.03, 1.05] | <0.01 | <0.001 | 1.02<br>[1.00, 1.04] | 0.01 | 0.04 |
|  | **Adulthood Carstairs deprivation**<br>(1 decile more deprived) | 1.06<br>[1.05, 1.07] | 0.01 | <0.001 | 1.04<br>[1.03, 1.06] | 0.01 | <0.001 |
| **Self-harm > Suicide**<br>(N = 13/516) | **Childhood Cognitive ability**<br>(1SD higher) | 1.11<br>[0.63, 1.96] | 0.29 | 0.71 | 1.08<br>[0.55, 2.14] | 0.35 | 0.82 |
|  | **Childhood Family position**<br>(1 person later) | 0.94<br>[0.54, 1.63] | 0.28 | 0.83 | 0.65<br>[0.30, 1.43] | 0.40 | 0.28 |
|  | **Childhood Family size**<br>(1 person larger) | 1.08<br>[0.86, 1.35] | 0.12 | 0.51 | 1.31<br>[0.91, 1.87] | 0.18 | 0.14 |
|  | **Adulthood Carstairs deprivation**<br>(1 decile more deprived) | 0.82<br>[0.68, 0.99] | 0.10 | 0.04 | 0.87<br>[0.71, 1.07] | 0.11 | 0.18 |
|  | **Time in self-harm state**<br>(1 month longer) | - | - | - | 1.04<br>[1.02, 1.06] | 0.01 | <0.001 |
| **Self-harm > Non-suicide death**<br>(N = 350/516) | **Childhood Cognitive ability**<br>(1SD higher) | 0.86<br>[0.77, 0.96] | 0.06 | 0.01 | 0.88<br>[0.78, 1.00] | 0.07 | 0.05 |
|  | **Childhood Family position**<br>(1 person later) | 1.07<br>[0.96, 1.20] | 0.05 | 0.19 | 1.00<br>[0.85, 1.19] | 0.09 | 0.96 |
|  | **Childhood Family size**<br>(1 person larger) | 1.03<br>[0.99, 1.08] | 0.02 | 0.16 | 1.01<br>[0.94, 1.09] | 0.04 | 0.76 |
|  | **Adulthood Carstairs deprivation**<br>(1 decile more deprived) | 1.04<br>[1.00, 1.08] | 0.02 | 0.05 | 1.05<br>[1.00, 1.10] | 0.02 | 0.03 |
|  | **Time in self-harm state**<br>(1 month longer) | - | - | - | 1.00<br>[1.00, 1.01] | <0.01 | <0.001 |

### Suicide risk

Among both males and females, the risk of transitioning from unaffected to suicide was not significantly associated with childhood cognitive ability or other covariates in either the univariate or fully-adjusted models.

Risk of transition from self-harm to suicide was not significantly associated with childhood cognitive ability in either the univariate or mutually-adjusted model, among either males or females. In the mutually-adjusted model, time spent in the self-harm state (before transition or censor) was significantly associated with the risk of suicide, with a single month increase in time associated with a 1% increase in suicide risk among males (HR = 1.01, 95% CI [1.00, 1.02]) and a 4% higher risk of suicide among females (HR = 1.01, 95% CI [1.00, 1.02]). For males only, an increased risk of suicide was also significantly associated with being born later in the family in the mutually-adjusted model. A 1-later birth position (e.g., second child vs. first child) was associated with a 189% higher risk (HR = 2.89, 95% CI [1.13, 7.40]).

### Non-suicide death risk

As in previous work in the same cohort (Calvin et al., 2017), an increased risk of transitioning from unaffected to non-suicide deaths was significantly associated with lower childhood cognitive ability, both in the univariate and fully-adjusted models and among males and females. In the fully-adjusted model, a 1SD advantage in childhood cognitive ability was associated with a 5% lower risk of death among males (HR = 0.95, 95% CI [0.92, 0.98]), and a 16% lower risk of death among females (HR = 0.84, 95% CI [0.81, 0.87]). The risk of transitioning from unaffected to non-suicide death was also significantly associated with area deprivation in adulthood; a single decile increase on the Carstairs index of deprivation was associated with a 6% higher risk of death among males (HR = 1.06, 95% CI [1.05, 1.07]) and a 4% higher risk of death among females (HR = 1.04, 95% CI [1.03, 1.06]). Among females only, a higher risk of transitioning from unaffected to non-suicide death was also significantly associated with larger family size in the fully-adjusted model; a 1-person increase in family size was associated with a 2% higher risk of death (HR = 1.02, 95% CI [1.00, 1.04]).

Among both males and females, an increased risk of transitioning from self-harm to non-suicide death was significantly associated with lower childhood cognitive ability, though only in the univariate model. Among females, only adulthood deprivation and time spent in the self-harm state were significantly associated with risk of transitioning from self-harm to non-suicide death in the mutually-adjusted model. A single decile increase in Carstairs deprivation index was associated with a 5% higher risk of death (HR = 1.05, 95% CI [1.00, 1.10]), and a single month increase in time associated with a <1% increase in risk (HR = 1.00, 95% CI [1.00, 1.01]). Among males, none of the variables – including time spent in the self-harm state – were significantly associated with risk of non-suicide death in the mutually-adjusted model.

## Discussion

Using a large population cohort, the present study investigated the association between childhood cognitive ability and both self-harm and suicide in later life. Higher cognitive ability in childhood was associated with reduced risk of self-harm and non-suicide death across later-life, but not with risk of suicide (regardless of whether individuals had experienced self-harm). Meanwhile, covariates including higher area-level deprivation and longer time spent living with self-harm showed consistent independent associations with higher risk of self-harm and suicide, respectively, among both males and females.

### Self-harm

Transitions into a self-harm state were relatively rare, particularly among males. Lower cognitive ability in childhood was associated with an increased risk of self-harm among unaffected individuals. Although this association was only significant among older females (HR = 0.90), a similar strength of association was noted among older males (HR = 0.90). This is broadly consistent with observations of an increased risk of various later-life mental health problems that are associated with self-harm, including depression, psychological distress and poor wellbeing, among males and females with lower cognitive ability in childhood (Bridger & Daly, 2017; Cheng & Furnham, 2013; Hatch et al., 2007; Iveson et al., 2020, 2021). The observed association is notably weaker than those reported in previous studies. For example, in a study of Swedish conscripted males, Batty et al. reported a significant and much stronger association between higher cognitive ability and lower risk of self-harm, even after accounting for similar covariates (HR = 0.64) (Batty et al., 2010). Note, however, that the follow-up period in Batty et al. primarily covered mid-life, where self-harm and suicide is more common than in later-life. The relative rarity of self-harm among older males in the present sample may have limited our statistical power. At the same time, the impact of cognitive ability on self-harm risk may dissipate over time as other factors become more important, resulting in weaker associations among older adults. Indeed, the present study observed a strong association between higher levels of poverty in adulthood and greater risk of self-harm in later life, along with an attenuation of the association between childhood cognitive ability and self-harm risk once socioeconomic conditions were taken into account. The importance of socioeconomic conditions for self-harm risk is consistent with work that used routinely-collected health data to track self-harm longitudinally (Carr et al., 2016; Kivimäki et al., 2020). Furthermore, hospitalisation due to self-harm has been highlighted as a starting point for the accumulation of further socioeconomically-patterned comorbidities, including liver disease, heart disease and dementia (Kivimäki et al., 2020).

### Suicide

One advantage of the multistate model used in the present study was its ability to separate the suicide risk of older adults experiencing a self-harm event during follow-up from that of unaffected individuals. We observed a higher risk of suicide among older adults experiencing self-harm, consistent with a great deal of work highlighting self-harm as a major risk factor for suicide (Hawton et al., 2003; Morgan et al., 2018; Murphy et al., 2012). However, this self-harm-related increase in risk was particularly high among females. Given that suicide was more frequent among older males, and that self-harm events represent hospital admissions, this likely indicates less help-seeking behaviour among males (De Leo et al., 2005) rather than sex differences in self-harm per se.

Despite separate estimation of suicide risk, childhood cognitive ability was not significantly associated with transitions to suicide from either the self-harm state or the unaffected state. This contrasts to work examining suicide in the same sample, in which lower childhood cognitive ability was associated with an increased risk of suicide among males but not females (Calvin et al., 2017). Note, however, that this association was not adjusted for socioeconomic circumstances. In the present study, there was evidence of confounding by socioeconomic circumstances; the marginally-significant association between lower childhood cognitive ability and higher suicide risk was attenuated once childhood and adulthood socioeconomic circumstances were accounted for. Meanwhile, Gunnell et al. report an association between lower cognitive ability (in early adulthood) and suicide risk that is only slightly attenuated when controlling for educational attainment (Gunnell et al., 2005). Although this may represent differences in the confounding contributions of education versus socioeconomic conditions (e.g., deprivation) or the impact of later assessment of cognitive ability, it more likely reflects substantial differences in the frequency of suicide events and the resulting impact on statistical power. In the present study, suicide was rare, affecting only 134 males and 44 females. Although the rate of suicides (0.4% of males and 0.1% of females) is similar to that observed by Gunnell et al. (0.2% of males), there are much fewer suicide events in the present study making it difficult to detect small but meaningful associations between cognitive ability and suicide.

Although cognitive ability was not significantly associated with suicide risk, one covariate was significantly associated with suicide risk among both males and females: time spent in the self-harm state. The risk of suicide was higher among those who spent longer in the self-harm state – i.e., among those who were further away in time from their self-harm hospitalisation. This may present a possible point for intervention to reduce suicide risk. However, it is unclear whether this represents the timing of the exposure, with earlier self-harm events posing the greatest risk, or the potential for repeated exposure, with those exposed early having the largest time window to experience self-harm again. Note that the present study does not account for the number of self-harm events accumulated across follow-up. It may be that older adults spending longer in the self-harm state experience repeated self-harm events and that these increase the risk of suicide (Murphy et al., 2012). Further work should aim to distinguish the impact of delaying self-harm onset versus reducing repeated self-harm events.

### Non-suicide death

Whereas suicide was a rare event, non-suicide death was relatively frequent, affecting over half of the sample over follow-up. Among unaffected individuals, lower childhood cognitive ability and higher adulthood deprivation were both significantly associated with increased risk of non-suicide death. Similar associations among unaffected individuals have been reported previously, both in the same sample (Calvin et al., 2017; Čukić et al., 2017; Iveson et al., 2018) and in others (Calvin et al., 2011; Christensen et al., 2016), and is consistent in its direction and strength.

Non-suicide death was particularly frequent among those experiencing self-harm, affecting around 70% of individuals. This is consistent with work showing significantly reduced life expectancy among those experiencing self-harm (Bergen et al., 2012). Indeed, work has shown that older adults experiencing self-harm exhibit increased risk of both suicide and non-suicide death within the year following self-harm, relative to age- and sex-matched controls without experience of self-harm (Morgan et al., 2018). Note, however, that in the present study, we observed a very weak association between longer time spent in the self-harm state and mortality risk among older adults experiencing self-harm, and only among females. Indeed, none of the included factors were significantly associated with non-suicide mortality risk among those experiencing self-harm, unlike among unaffected individuals. Increased non-suicide mortality risk among older adults experiencing self-harm may instead result from factors not captured in the present study, including greater multimorbidity and polypharmacy (Mitchell et al., 2017; Morgan et al., 2018).

### Potential mechanisms

A greater susceptibility to self-harm among those with lower childhood cognitive ability may arise due to indirect routes, mediated by factors arising between childhood and later-life. For example, childhood cognitive ability has been associated with employment outcomes and quality of life (Iveson & Deary, 2019) and with health behaviours (Fawns-Ritchie et al., 2018). These factors may in turn increase the risk of self-harm in later life. Lower childhood cognitive ability may also indirectly impact self-harm risk through its contribution to insufficient coping strategies (Kitano & Lewis, 2005). The impact of lower childhood cognitive ability may also be through a more direct route. Note that this is not mutually exclusive with more indirect mechanisms. The system integrity theory (Deary, 2012) suggests that cognitive ability is an indicator of bodily functioning, something that is supported by genetic studies of cognitive ability and many health outcomes (Arden et al., 2016). Similarly, Gunnell et al. suggest that lower cognitive test scores in early-life may indicate impaired neurodevelopment, including in those regions of the brain implicated in psychiatric illness (Gunnell et al., 2005). For example, lower childhood cognitive ability has been associated with higher depression risk (Iveson et al., 2021; van Os, 1997), a condition that greatly increases the risk of self-harm among older adults (J. Chan et al., 2007; Dennis et al., 2005; Draper, 1996; Morgan et al., 2018; Szanto et al., 2002). However, psychiatric illness is unlikely to explain all of cognitive ability’s association with suicide risk; work has noted that cognitive ability better predicts self-harm risk among those without psychosis than those with psychosis (Batty et al., 2010).

### Strengths & Limitations

This study builds on previous work by extending follow-up further into later-life, up to age 85 years-old, and by modelling independent associations with suicide risk among those with and without self-harm events. Furthermore, by focussing on cognitive ability in childhood – decades before the observation of self-harm and suicide – the present study better addresses concerns over confounding. However, the present study is subject to several important limitations. As with many health record linkage studies, the present study does not include self-harm events that do not result in a hospital admission. This misses self-harm events that present only to primary care or emergency departments (without subsequent admission) and those that do not present to healthcare services at all, likely underestimating potential risk factors (Marchant et al., 2020). Furthermore, individuals in the sample are treated as ‘healthy’ at entry to the study; no information is available regarding the history of self-harm or existing psychiatric illness prior to the start of the study. Work has highlighted the importance of previous self-harm and previous psychiatric illness for self-harm risk in later life (Troya et al., 2019). Similarly, the present study does not distinguish the intention behind self-harm events. Although all included events were deliberate, it is unclear whether they were attempts at suicide or not. Work has shown that attempted suicide makes up a large proportion of self-harm events in older adults (Draper, 1996; Hawton & Harriss, 2008). Self-harm events in the present sample, then, are more likely to represent suicide attempts, particularly since they require hospitalisation. Finally, the definition for suicide events used in the present study differs to many others by only including deaths due to intentional self-harm, not deaths of undetermined intent. In the present study, restricting the suicide definition likely underestimates the contribution of childhood cognitive ability, particularly given the rarity of suicide events. Indeed, the increased case numbers captured by a wider criteria appears to narrow confidence intervals while maintaining cognitive ability effect sizes (Calvin et al., 2017).

Perhaps most notably, the rarity of self-harm and suicide events in the present study poses a challenge in terms of the statistical power to detect meaningful associations, one that has been highlighted by several similar studies (Hawton et al., 2003; Murphy et al., 2012). Much larger samples, such as the population-wide cohorts used in conscript studies (N = 987,308 males) (Gunnell et al., 2005), are needed to investigate these rare events. Note, however, that existing population-wide cohorts are themselves limited in their focus (e.g., males only); new longitudinal cohorts of unprecedented scale and follow-up are needed to adequately address questions about early-life risk factors for suicide.

## Conclusions

Self-harm and suicide remain prevalent among older adults and, given the ageing population, present major challenges to public health. The present study adds to the growing literature regarding the life-long consequences of childhood disadvantage and supports the need for early-life interventions to improve health across the life course. It further demonstrates that the association between lower childhood cognitive ability and higher self-harm risk is similar between males and females, but highlights the relative rarity of self-harm events among older adults, at least as captured by hospital admissions. It also demonstrates that the impact of lower childhood cognitive ability differs between self-harm and suicide risk. Indeed, the present study shows no significant association with suicide risk when modelled concurrently with self-harm. This suggests that tackling early-life inequalities (including in cognitive ability, through education etc.) may reduce the risk of self-harm, but may only indirectly affect suicide risk through self-harm prevention. Reducing suicide risk more specifically may be better achieved at the primary care level, through screening and management programmes (Okolie et al., 2017). Further work is required to clarify the mechanisms underlying associations with self-harm, and to re-examine such rare events in much larger cohorts.

## Data Availability

All data produced in the present study are available upon ethical approval, public benefit approval (through the Public Benefit and Privacy Panel) and reasonable request to the data controllers.

## References

Allebeck, P., Allgulander, C., & Fisher, L. D. (1988). Predictors of completed suicide in a cohort of 50,465 young men: Role of personality and deviant behaviour. BMJ, 297(6642), 176–178. https://doi.org/10.1136/bmj.297.6642.176

Andersson, L., Allebeck, P., Gustafsson, J.-E., & Gunnell, D. (2008). Association of IQ scores and school achievement with suicide in a 40-year follow-up of a Swedish cohort. Acta Psychiatrica Scandinavica, 118(2), 99–105. https://doi.org/10.1111/j.1600-0447.2008.01171.x

Arden, R., Luciano, M., Deary, I. J., Reynolds, C. A., Pedersen, N. L., Plassman, B. L., McGue, M., Christensen, K., & Visscher, P. M. (2016). The association between intelligence and lifespan is mostly genetic. International Journal of Epidemiology, 45(1), 178–185. https://doi.org/10.1093/ije/dyv112

Batty, G. D., Kivimäki, M., Bell, S., Gale, C. R., Shipley, M., Whitley, E., & Gunnell, D. (2018). Psychosocial characteristics as potential predictors of suicide in adults: An overview of the evidence with new results from prospective cohort studies. Translational Psychiatry, 8(1), 22. https://doi.org/10.1038/s41398-017-0072-8

Batty, G. D., Whitley, E., Deary, I. J., Gale, C. R., Tynelius, P., & Rasmussen, F. (2010). Psychosis alters association between IQ and future risk of attempted suicide: Cohort study of 1 109 475 Swedish men. BMJ, 340(jun03 1), c2506–c2506. https://doi.org/10.1136/bmj.c2506

Bergen, H., Hawton, K., Waters, K., Ness, J., Cooper, J., Steeg, S., & Kapur, N. (2012). Premature death after self-harm: A multicentre cohort study. The Lancet, 380(9853), 1568–1574. https://doi.org/10.1016/S0140-6736(12)61141-6

Bridger, E., & Daly, M. (2017). Does Cognitive Ability Buffer the Link Between Childhood Disadvantage and Adult Health? Health Psychology, 36(10), 966–976. https://doi.org/10.1037/hea0000538

Calvin, C. M., Batty, G. D., Der, G., Brett, C. E., Taylor, A., Pattie, A., Cukic, I., & Deary, I. J. (2017). Childhood intelligence in relation to major causes of death in 68 year follow-up: Prospective population study. BMJ (Online), 357. https://doi.org/10.1136/bmj.j2708

Calvin, C. M., Deary, I. J., Fenton, C., Roberts, B. A., Der, G., Leckenby, N., & Batty, G. D. (2011). Intelligence in youth and all-cause-mortality: Systematic review with meta-analysis. International Journal of Epidemiology, 40(3), 626–644. https://doi.org/10.1093/ije/dyq190

Carr, M. J., Ashcroft, D. M., Kontopantelis, E., Awenat, Y., Cooper, J., Chew-Graham, C., Kapur, N., & Webb, R. T. (2016). The epidemiology of self-harm in a UK-wide primary care patient cohort, 2001–2013. BMC Psychiatry, 16(1), 53. https://doi.org/10.1186/s12888-016-0753-5

Carstairs, V., & Morris, R. (1991). Deprivation and Health in Scotland. Aberdeen University Press.

Chan, J., Draper, B., & Banerjee, S. (2007). Deliberate self-harm in older adults: A review of the literature from 1995 to 2004. International Journal of Geriatric Psychiatry, 22(8), 720–732. https://doi.org/10.1002/gps.1739

Chan, M. K. Y., Bhatti, H., Meader, N., Stockton, S., Evans, J., O’Connor, R. C., Kapur, N., & Kendall, T. (2016). Predicting suicide following self-harm: Systematic review of risk factors and risk scales. British Journal of Psychiatry, 209(4), 277–283. https://doi.org/10.1192/bjp.bp.115.170050

Cheng, H., & Furnham, A. (2013). Factors Influencing Adult Physical Health after Controlling for Current Health Conditions: Evidence from a British Cohort. PLoS ONE, 8(6), e66204. https://doi.org/10.1371/journal.pone.0066204

Christensen, G. T., Mortensen, E. L., Christensen, K., & Osler, M. (2016). Intelligence in young adulthood and cause-specific mortality in the Danish Conscription Database – A cohort study of 728,160 men. Intelligence, 59, 64–71. https://doi.org/10.1016/j.intell.2016.08.001

Čukić, I., Brett, C. E., Calvin, C. M., Batty, G. D., & Deary, I. J. (2017). Childhood IQ and survival to 79: Follow-up of 94% of the Scottish Mental Survey 1947. Intelligence, 63(May), 45–50. https://doi.org/10.1016/j.intell.2017.05.002

De Leo, D., Cerin, E., Spathonis, K., & Burgis, S. (2005). Lifetime risk of suicide ideation and attempts in an Australian community: Prevalence, suicidal process, and help-seeking behaviour. Journal of Affective Disorders, 86(2–3), 215–224. https://doi.org/10.1016/j.jad.2005.02.001

de Wreede, L. C. de, Fiocco, M., & Putter, H. (2011). mstate: An R Package for the Analysis of Competing Risks and Multi-State Models. Journal of Statistical Software, 38(7). https://doi.org/10.18637/jss.v038.i07

Deary, I. J. (2012). Looking for ‘System Integrity’ in Cognitive Epidemiology. Gerontology, 58(6), 545–553. https://doi.org/10.1159/000341157

Dennis, M., Wakefield, P., Molloy, C., Andrews, H., & Friedman, T. (2005). Self-harm in older people with depression: Comparison of social factors, life events and symptoms. British Journal of Psychiatry, 186(6), 538–539. https://doi.org/10.1192/bjp.186.6.538

Draper, B. (1996). Attempted suicide in old age. International Journal of Geriatric Psychiatry, 11(7), 577–587. https://psycnet.apa.org/doi/10.1002/(SICI)1099-1166(199607)11:7%3C577::AID-GPS362%3E3.0.CO;2-V

Duarte, T. A., Paulino, S., Almeida, C., Gomes, H. S., Santos, N., & Gouveia-Pereira, M. (2020). Self-harm as a predisposition for suicide attempts: A study of adolescents’ deliberate self-harm, suicidal ideation, and suicide attempts. Psychiatry Research, 287, 112553. https://doi.org/10.1016/j.psychres.2019.112553

Fawns-Ritchie, C., Starr, J. M., & Deary, I. J. (2018). Role of cognitive ability in the association between functional health literacy and mortality in the Lothian Birth Cohort 1936: A prospective cohort study. BMJ Open, 8(9), e022502. https://doi.org/10.1136/bmjopen-2018-022502

Fergusson, D. M., Horwood, L. J., & Ridder, E. M. (2005). Show me the child at seven II: Childhood intelligence and later outcomes in adolescence and young adulthood. Journal of Child Psychology and Psychiatry and Allied Disciplines, 46(8), 850–858. https://doi.org/10.1111/j.1469-7610.2005.01472.x

Gunnell, D., Magnusson, P. K. E., & Rasmussen, F. (2005). Low intelligence test scores in 18 year old men and risk of suicide: Cohort study. BMJ, 330(7484), 167. https://doi.org/10.1136/bmj.38310.473565.8F

Hatch, S. L., Jones, P. B., Kuh, D., Hardy, R., Wadsworth, M. E. J., & Richards, M. (2007). Childhood cognitive ability and adult mental health in the British 1946 birth cohort. Social Science and Medicine, 64(11), 2285–2296. https://doi.org/10.1016/j.socscimed.2007.02.027

Hawton, K., & Harriss, L. (2008). How Often Does Deliberate Self-Harm Occur Relative to Each Suicide? A Study of Variations by Gender and Age. Suicide and Life-Threatening Behavior, 38(6), 650–660. https://doi.org/10.1521/suli.2008.38.6.650

Hawton, K., Zahl, D., & Weatherall, R. (2003). Suicide following deliberate self-harm: Long-term follow-up of patients who presented to a general hospital. British Journal of Psychiatry, 182(6), 537–542. https://doi.org/10.1192/bjp.182.6.537

Hemmingsson, T., Melin, B., Allebeck, P., & Lundberg, I. (2006). The association between cognitive ability measured at ages 18–20 and mortality during 30 years of follow-up— A prospective observational study among Swedish males born 1949–51. International Journal of Epidemiology, 35(3), 665–670. https://doi.org/10.1093/ije/dyi321

Iveson, M. H., Čukič, I., Der, G., Batty, G. D., & Deary, I. J. (2018). Intelligence and all-cause mortality in the 6-Day Sample of the Scottish Mental Survey 1947 and their siblings: Testing the contribution of family background. International Journal of Epidemiology, 47(1). https://doi.org/10.1093/ije/dyx168

Iveson, M. H., & Deary, I. J. (2019). Early-life predictors of retirement decisions and post-retirement health. SSM - Population Health, 8. https://doi.org/10.1016/j.ssmph.2019.100430

Iveson, M. H., Dibben, C., & Deary, I. J. (2020). Early-life circumstances and the risk of function-limiting long-term conditions in later life. Longitudinal and Life Course Studies, 11(2), 157–180. https://doi.org/10.1332/175795919X15762565000695

Iveson, M. H., Taylor, A., Harris, S. E., Deary, I. J., & McIntosh, A. M. (2021). Apolipoprotein E e4 allele status and later-life depression in the Lothian Birth Cohort 1936. Psychological Medicine, 1–9. https://doi.org/10.1017/S0033291721000623

Jiang, G.-X., Rasmussen, F., & Wasserman, D. (1999). Short stature and poor psychological performance: Risk factors for attempted suicide among Swedish male conscripts. Acta Psychiatrica Scandinavica, 100(6), 433–440. https://doi.org/10.1111/j.1600-0447.1999.tb10893.x

Kitano, M. K., & Lewis, R. B. (2005). Resilience and coping: Implications for gifted children and youth at risk. Roeper Review, 27(4), 200–205. https://doi.org/10.1080/02783190509554319

Kivimäki, M., Batty, G. D., Pentti, J., Shipley, M. J., Sipilä, P. N., Nyberg, S. T., Suominen, S. B., Oksanen, T., Stenholm, S., Virtanen, M., Marmot, M. G., Singh-Manoux, A., Brunner, E. J., Lindbohm, J. V., Ferrie, J. E., & Vahtera, J. (2020). Association between socioeconomic status and the development of mental and physical health conditions in adulthood: A multi-cohort study. The Lancet Public Health, 5(3), e140–e149. https://doi.org/10.1016/S2468-2667(19)30248-8

Kuh, D., Richards, M., Hardy, R., Butterworth, S., & Wadsworth, M. E. J. (2004). Childhood cognitive ability and deaths up until middle age: A post-war birth cohort study. International Journal of Epidemiology, 33(2), 408–413. https://doi.org/10.1093/ije/dyh043

Linsley, K. R., Schapira, K., & Kelly, T. P. (2001). Open verdict v. Suicide – importance to research. British Journal of Psychiatry, 178(5), 465–468. https://doi.org/10.1192/bjp.178.5.465

Marchant, A., Turner, S., Balbuena, L., Peters, E., Williams, D., Lloyd, K., Lyons, R., & John, A. (2020). Self-harm presentation across healthcare settings by sex in young people: An e-cohort study using routinely collected linked healthcare data in Wales, UK. Archives of Disease in Childhood, 105(4), 347–354. https://doi.org/10.1136/archdischild-2019-317248

McLoone, P. (2004). Carstairs scores for Scottish postcode sectors from the 2001 Census. MRC Social and Public Health Sciences Unit.

Mitchell, R., Draper, B., Harvey, L., Brodaty, H., & Close, J. (2017). The association of physical illness and self-harm resulting in hospitalisation among older people in a population-based study. Aging & Mental Health, 21(3), 279–288. https://doi.org/10.1080/13607863.2015.1099610

Morgan, C., Webb, R. T., Carr, M. J., Kontopantelis, E., Chew-Graham, C. A., Kapur, N., & Ashcroft, D. M. (2018). Self-harm in a primary care cohort of older people: Incidence, clinical management, and risk of suicide and other causes of death. The Lancet Psychiatry, 5(11), 905–912. https://doi.org/10.1016/S2215-0366(18)30348-1

Murphy, E., Kapur, N., Webb, R., Purandare, N., Hawton, K., Bergen, H., Waters, K., & Cooper, J. (2012). Risk factors for repetition and suicide following self-harm in older adults: Multicentre cohort study. British Journal of Psychiatry, 200(5), 399–404. https://doi.org/10.1192/bjp.bp.111.094177

Nathanson, C. A. (1984). Sex Differences in Mortality. Annual Review of Sociology, 10(1), 191–213. https://doi.org/10.1146/annurev.so.10.080184.001203

Okolie, C., Dennis, M., Simon Thomas, E., & John, A. (2017). A systematic review of interventions to prevent suicidal behaviors and reduce suicidal ideation in older people. International Psychogeriatrics, 29(11), 1801–1824. https://doi.org/10.1017/S1041610217001430

ONS. (2021). Suicides in England and Wales: 2020 registrations. Office for National Statistics. https://www.ons.gov.uk/peoplepopulationandcommunity/birthsdeathsandmarriages/deaths/bulletins/suicidesintheunitedkingdom/2020registrations

Osler, M., Nybo Andersen, A.-M., & Nordentoft, M. (2008). Impaired childhood development and suicidal behaviour in a cohort of Danish men born in 1953. Journal of Epidemiology & Community Health, 62(1), 23–28. https://doi.org/10.1136/jech.2006.053330

O’Toole, B. I., & Cantor, C. (1995). Suicide risk factors among Australian Vietnam era draftees. Suicide & Life-Threatening Behavior, 25(4), 475–488.

R Core Team. (2021). R: A Language and environment for statistical computing. R Foundation for Statistical Computing. https://www.r-project.org/

Read, J. G., & Gorman, B. K. (2010). Gender and Health Inequality. Annual Review of Sociology, 36(1), 371–386. https://doi.org/10.1146/annurev.soc.012809.102535

Scottish Public Health Observatory. (2022). Suicide: Scottish trends. Public Health Scotland. https://www.scotpho.org.uk/health-wellbeing-and-disease/suicide/data/scottish-trends/

Szanto, K., Gildengers, A., Mulsant, B. H., Brown, G., Alexopoulos, G. S., & Reynolds, C. F. (2002). Identification of Suicidal Ideation and Prevention of Suicidal Behaviour in the Elderly: Drugs & Aging, 19(1), 11–24. https://doi.org/10.2165/00002512-200219010-00002

The Scottish Council for Research in Education. (1949). The trend of Scottish intelligence. University of London Press.

Troya, M. I., Babatunde, O., Polidano, K., Bartlam, B., McCloskey, E., Dikomitis, L., & Chew-Graham, C. A. (2019). Self-harm in older adults: Systematic review. British Journal of Psychiatry, 214(4), 186–200. https://doi.org/10.1192/bjp.2019.11

van Os, J. (1997). Developmental Precursors of Affective Illness in a General Population Birth Cohort. Archives of General Psychiatry, 54(7), 625. https://doi.org/10.1001/archpsyc.1997.01830190049005

